# Bayesian Spatiotemporal Small Area Estimation of HIV Testing Uptake in Ghana, 2008 to 2022: Combining Machine Learning Geospatial Covariates with District Level BYM2 and RW1 Modelling of the Ghana Demographic and Health Surveys

**DOI:** 10.64898/2026.07.31.26359204

**Authors:** Osman Abdul-Fatawu Iddrisu, Abubakar Hudu Siddick, Abubakar Iddrisu Siddiq

## Abstract

**Background:** HIV testing is the entry point into the diagnosis, treatment and viral suppression cascade, yet in many low- and middle-income countries the household surveys used to monitor testing coverage are not powered for estimation below the regional level. We produced calibrated district level estimates of HIV testing uptake among women in Ghana across three Demographic and Health Survey (DHS) rounds and examined the spatial and temporal structure of the heterogeneity that remained once measured covariates were accounted for.

**Methods:** We pooled individual recode and geo referenced cluster data from the 2008, 2014 and 2022 Ghana DHS (n = 4,769, 9,391 and 15,014 women respectively; outcome: ever tested for HIV, variable v781), aggregated to 261 level two administrative districts by survey round, and fitted a Bayesian hierarchical binomial model that combined a BYM2 conditional autoregressive spatial term, a first order random walk (RW1) temporal term, and three standardized covariates: WorldPop population density derived from a machine learning dasymetric algorithm, Malaria Atlas Project travel time to the nearest city, and cluster urban proportion, each extracted within buffers around cluster coordinates that matched the DHS displacement protocol. Inference used integrated nested Laplace approximation (INLA) implemented through R-INLA (Lindgren and Rue, 2015). Residual spatial structure was assessed with global and local Moran’s I.

**Results:** National crude testing prevalence rose from 20.7% in 2008 to 46.7% in 2014 and 53.8% in 2022. District sample sizes were small and unevenly distributed (2008 median n = 20 women per district; 91.8% of districts had fewer than 50), which is why model based smoothing rather than direct estimation was required. Urban cluster proportion was independently associated with higher testing odds (odds ratio [OR] 1.10, 95% credible interval [CrI] 1.04 to 1.16 per one standard deviation increase) and travel time to the nearest city with lower odds (OR 0.90, 95% CrI 0.84 to 0.96); population density showed no independent association once these two variables were included (OR 0.95, 95% CrI 0.89 to 1.02). The spatial mixing parameter of the BYM2 term (phi = 0.716, 95% CrI 0.498 to 0.887) indicated that around seven tenths of spatially attributable variance was structured rather than idiosyncratic. Global Moran’s I on the fitted spatial effect surface was 0.616 (p = 6.6 x 10 to the power minus 60), and local indicators of spatial association identified a contiguous low uptake cluster across the northern regions together with three compact high uptake clusters in the south-central corridor.

**Conclusions:** Combining machine learning derived geospatial covariates with an explicit spatiotemporal Bayesian hierarchy exposes a persistent north to south gradient in HIV testing uptake that measured accessibility and urbanicity do not fully explain and identifies specific district clusters as priorities for targeted testing scale up.

## 1. Introduction

HIV testing is the entry point into the diagnosis, treatment and viral suppression cascade that anchors current global HIV control targets. A person who does not know their status cannot be linked to antiretroviral therapy, and population level progress toward epidemic control depends on testing coverage that is both high and geographically even. In practice it is rarely either. National household surveys, and above all the Demographic and Health Surveys, remain the principal instrument used to track self-reported testing uptake in countries such as Ghana, but they are designed to yield precise estimates at the national and regional level rather than at the district level where testing programmes are actually planned, staffed and resourced.

A substantial body of Ghana specific survey research has documented who does and does not get tested, and why. (1) applied Anderson’s behavioural model of health service use to explain testing uptake among sexually active men, and (2) reported similarly patterned determinants of testing among Ghanaian men using nationally representative data. (3) showed that rural residence remained a persistent correlate of lower testing uptake among sexually active Ghanaians even after adjustment for individual level covariates, a finding echoed by (4) in an analysis restricted to women of reproductive age, and by (5) in an earlier national assessment of testing decisions. (6) and (7) narrowed the same question to specific sub populations, tertiary students in Hohoe and youth in Kintampo South District respectively and found uptake patterns that diverged sharply from the national average in ways that a single national statistic cannot reveal. More recently, (8) and (9) used the 2022 GDHS to examine testing uptake and self-test awareness among Ghanaian men and women respectively, (10) analyzed correlates of testing among men in the same survey round, and (11) decomposed trends and inequalities in antenatal HIV testing across exactly the 2008 to 2022 window examined here. (12) focused on young women, and (13) used the Multiple Indicator Cluster Survey to extend the evidence base on men beyond the DHS platform itself.

None of these studies, individually or collectively, resolves the problem that motivates the present analysis. Every one of them models testing uptake as a function of individual and household characteristics summarized at the regional or national level, and none produces a district level estimate with an attached measure of uncertainty. (14) came closest, mapping district level HIV testing prevalence from the 2014 GDHS using design based methods and a complex survey analysis and identified marked regional disparities that recur throughout the literature just reviewed. (15) took a complementary approach, mapping geographical access to point of care diagnostic testing, including HIV testing, within the Bono Region specifically, and (16) examined the spatial distribution of women’s health screening behaviour more broadly. These mapping studies share two limitations. First, each analyses a single survey round in isolation, so a district’s estimate in one year borrows no information from its own trajectory in adjacent rounds, and genuine change over time cannot be distinguished from sampling variation between independent samples. Second, the district sample sizes underlying a single round of the GDHS are frequently too small to support a stable direct estimate at all, a problem that design based analysis of one round cannot itself correct.

Two adjacent bodies of work suggest how both limitations can be addressed at once. Wakefield and colleagues formalized spatiotemporal small area estimation frameworks that borrow strength jointly across geography and survey round while propagating uncertainty coherently (17), an approach since adapted to HIV burden estimation in Nigeria by (18), in Zambia by (19), and combined with continent wide age and sex specific mapping by (20) and (21), while (22) built a comparable district level modelling tool, Naomi, specifically to support UNAIDS estimation of HIV indicators between survey rounds. Separately, geospatial covariates produced by machine learning pipelines, WorldPop’s random forest dasymetric population surfaces and comparable accessibility layers among them, have been shown to sharpen the covariate basis available to disease mapping models even where the outcome itself has nothing to do with the algorithm that generated the covariate. (23) went further still and embedded a graph neural network directly inside a Bayesian small area estimation framework with spatial regularisation, an architectural fusion of deep learning and hierarchical Bayesian inference rather than a covariate level one. (24) reviewed the broader convergence of artificial intelligence and infectious disease modelling in similar terms, and (25) documented the rapid growth of artificial intelligence applications specifically within HIV care, underscoring that the methodological ingredients this paper combines are each independently active areas of development.

No published analysis, to our knowledge, has combined a spatiotemporal Bayesian hierarchy spanning multiple GDHS rounds, geospatial covariates produced by machine learning methods, and formal residual spatial diagnostics, for HIV testing uptake in Ghana specifically. This paper closes that gap and, in doing so, addresses four questions. First, how has district level HIV testing uptake in Ghana evolved between 2008 and 2022 once sampling noise is separated from genuine geographic and temporal signal. Second, do population density and accessibility surfaces derived from machine learning methods carry independent explanatory power for testing uptake once conventional survey covariates are included. Third, after fitting a spatially and temporally smoothed model, does statistically detectable spatial clustering remain, and where is it located? Fourth, what do the resulting small area estimates imply for the geographic targeting of testing scale up resources in Ghana.

We argue that combining a BYM2 spatial term with a random walk temporal term and covariates derived from machine learning methods is not simply a matter of adding more predictors to an existing spatial model. The three elements address three distinct sources of the estimation problem. The BYM2 term borrows strength across geographically neighbouring districts to stabilize estimates built on small survey counts. The RW1 term borrows strength across survey rounds so that a district’s own history informs its current estimate, addressing the single round limitation of (14) and related mapping work. The machine learning derived covariates supply exposure information at a spatial resolution the survey sample cannot itself provide, particularly in low cluster density districts where too few DHS points exist to characterize local accessibility from the survey data alone. Used together within one coherent likelihood, these three components let the residual spatial and temporal random effects absorb only the structure that measured covariates cannot explain, which is what makes our residual diagnostic analysis informative rather than circular.

## 2. Methods

### 2.1 Data sources and study design

We used individual recode files from the 2008, 2014 and 2022 rounds of the Ghana Demographic and Health Survey (GHIR5AFL, GHIR72FL and GHIR8CFL), restricted to women with a non-missing response to variable v781, ever tested for HIV, which yielded analytic samples of 4,769, 9,391 and 15,014 women respectively. Each round was linked to its corresponding geo referenced cluster file containing displaced GPS coordinates for the primary sampling unit, following the DHS Program standard displacement of up to 2 km for urban clusters and 5 km for rural clusters, with 1% of rural clusters displaced up to 10 km; clusters with unset (0,0) coordinates were excluded. Clusters were aggregated to the number of women interviewed and the number reporting prior testing, then spatially joined to Ghana level two administrative boundaries (GADM v4) using coordinate reference system aligned point in polygon matching. Of 1,445 geo referenced clusters across the three rounds, 1,443 (99.9%) matched to a district; two clusters lying outside all polygons were dropped. Cluster counts per round were 404 in 2008, 423 in 2014 and 618 in 2022.

Cluster level counts were summed within district round cells to give the district round numerator, number tested, and denominator, number of women interviewed, that form the outcome of the Bayesian model. A full factorial grid of all districts by all three rounds was constructed so that district rounds without any interviewed cluster were represented as structurally missing rather than omitted, which is the correct treatment under a latent Gaussian field model: INLA integrates over the fitted linear predictor for these cells using the estimated spatial and temporal structure without contributing a likelihood term. The number of districts with at least one interviewed woman was 196 in 2008, 219 in 2014 and 237 in 2022, out of 261 districts on the analysis grid; the shortfall reflects the coverage of the DHS cluster sample rather than any missing data mechanism related to testing behaviour itself.

### 2.2 Outcome and small sample motivation for smoothing

Table 1 reports the crude, unsmoothed, national testing prevalence in each round, and Table 2 the distribution of women interviewed per district. The rise in median district sample size from 20 in 2008 to 33 in 2014 and 48 in 2022 reflects the expanding GDHS cluster sample over time, but even in 2022 more than half of districts, 52.7%, contributed fewer than 50 women. At n = 20 and an assumed true proportion near the observed national average of 0.21, the binomial standard error of a direct district estimate exceeds 9 percentage points, larger than the entire national increase in testing uptake between any two rounds. Direct estimates at this sample size are not usable for district level programme planning without some form of statistical smoothing, which is the quantitative justification specific to this dataset for the small area framework adopted below, and the reason a single round design based analysis of the kind reported by (14) is necessarily more precise in the regions with denser cluster coverage than in those with sparser coverage.

**Table 1.** National crude HIV testing prevalence among women, by GDHS round.

| Round | Districts with data | Women interviewed | Women tested | Crude prevalence |
| --- | --- | --- | --- | --- |
| 2008 | 196 | 4,669 | 966 | 20.7% |
| 2014 | 219 | 9,274 | 4,332 | 46.7% |
| 2022 | 237 | 15,014 | 8,078 | 53.8% |

**Table 2.** Distribution of district level sample sizes, women interviewed per district, by GDHS round.

| Round | Median n | IQR | % districts n<20 | % districts n<50 |
| --- | --- | --- | --- | --- |
| 2008 | 20 | 18.3 | 49.5% | 91.8% |
| 2014 | 33 | 31.0 | 12.8% | 72.6% |
| 2022 | 48 | 57.0 | 11.4% | 52.7% |

### 2.3 Geospatial covariates

Three continuous district round covariates were constructed from gridded surfaces external to the DHS instrument. Population density was extracted from WorldPop’s 100 m gridded population surfaces for the years matching each survey round, 2008, 2014 and 2020 as the closest available year to 2022. These surfaces are produced through a random forest based dasymetric redistribution of national census counts that uses ancillary land cover, settlement and infrastructure layers classified by machine learning methods, and we treat them here as a machine learning derived covariate rather than a raw census count. Travel time to the nearest population centre of 50,000 or more was extracted from the Malaria Atlas Project’s 2015 global friction surface accessibility layer, retrieved through the malariaAtlas R package, a cost distance surface built from a classified land cover and infrastructure friction map. An annual VIIRS night time lights composite was sought as a third proxy for local economic activity and urbanicity, but the Earth Observation Group repository returned authentication walled HTML responses rather than the requested raster files for all three years; this covariate was therefore dropped from the analysis rather than imputed, and we report its absence as a limitation in Section 4.6 rather than disguise the gap.

For each cluster, the relevant raster was averaged within a circular buffer around the cluster’s displaced coordinates, 2 km for clusters classified urban and 5 km for those classified rural in the corresponding DHS round, so that the buffer radius matched or exceeded the DHS Program’s own maximum plausible positional error. Cluster level covariate values were averaged, unweighted, within each district round to produce district round means, then centred and scaled to unit variance before inclusion as fixed effects. A fourth fixed effect, the proportion of a district round’s interviewed clusters classified as urban in the DHS sampling frame, was constructed directly from survey metadata and standardized in the same way.

### 2.4 Bayesian hierarchical spatiotemporal model

Let *i* = 1 to *I* index the *I* = 261 districts on the analysis grid and *t* = 1, 2, 3 index the three survey rounds, 2008, 2014 and 2022. Let *y*_it_ denote the number of women reporting prior HIV testing in district *i* at round *t*, and *n*_it_ the number of women interviewed in that district-round. We modelled

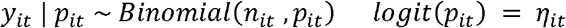

with linear predictor

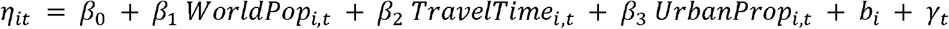

where all covariates are the standardised quantities defined in Section 2.3, *b*_i_ is a district-level spatial random effect common to all rounds, and _γt_ is a round-specific temporal random effect common to all districts. Fixed effects were assigned the software-default diffuse Gaussian prior,

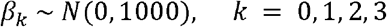

#### 2.4.1 BYM2 spatial term

The spatial effect followed the BYM2 formulation of Riebler, Sørbye, Simpson and Rue (2016), which re-expresses the classical convolution of an intrinsic conditional autoregressive component and an independent component as a single scaled mixture:

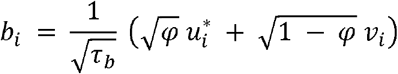

where *u*_i_ * is a variance-scaled intrinsic conditional autoregressive spatial component satisfying the improper pairwise-difference prior

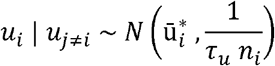

*n*_i_ is the number of neighbours of district *i* under Queen contiguity, *v*_i_ ∼ N(0,1) independently, _τb_ is the marginal precision of the combined effect, and _φc_ [0,1] is the proportion of marginal variance attributable to the spatially structured component. The adjacency structure was built with Queen contiguity polygon adjacency on the 261-district GADM level-two layer using the spdep package. Following current recommended practice, the intrinsic conditional autoregressive component was variance-scaled so that the geometric mean of the marginal variances across the graph equals 1, allowing _τb_ and _φ_ to be interpreted on a common, graph-independent scale. Penalised complexity priors were used throughout: _τb_ was assigned a penalized complexity prior with

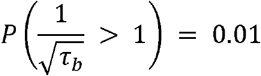

and _φ_ a penalized complexity prior with

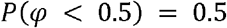

corresponding to a weakly informative baseline scepticism toward strong spatial structure that the data are free to overturn.

#### 2.4.2 Temporal term

The temporal effect _γt_ followed a first-order random walk over the ordinal round index *t* = 1, 2, 3:

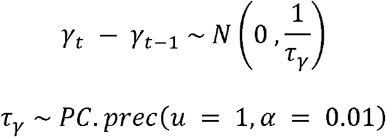

This choice treats the three rounds as equally spaced, which is an approximation given the true gaps of 6 years between 2008 and 2014 and 8 years between 2014 and 2022; we return to this in the limitations.

### 2.5 Position of machine learning components within the pipeline

We distinguish two ways that machine learning or deep learning methods can enter a small area estimation pipeline. The first is upstream, as the algorithm used to construct covariate surfaces that are then fed into an otherwise conventional Bayesian hierarchy. The second is architectural, where a learned component is embedded inside the latent field specification itself, as in the graph neural network small area estimator of (23), which combines spatial regularization, heterogeneous spatial units and Bayesian inference within a single trained architecture. The analysis reported here uses machine learning in the first sense only. The WorldPop population surface and the Malaria Atlas Project accessibility surface are themselves outputs of machine learning pipelines, random forest dasymetric mapping and classified friction cost distance modelling respectively, but the inferential layer that combines them with the survey data is a standard BYM2 and RW1 Bayesian hierarchy fitted by INLA, not a trained neural network. We consider this an important methodological boundary to state plainly rather than blur and return to an architecturally embedded alternative.

### 2.6 Residual spatial diagnostics

After model fitting, we extracted the posterior mean of the district level spatial effect from the INLA output for the BYM2 term. INLA’s internal representation of a bym2 latent component stores 2*I* values per posterior draw: the first *I* correspond to the combined effect *b_i_*, the quantity entering the linear predictor, and the second *I* to the intrinsic conditional autoregressive component alone. We indexed the first *I* values, so the quantity we term the structured spatial effect throughout the Results is the full combined BYM2 surface rather than the purely autoregressive component in isolation; because _φ_ was estimated at 0.716, the combined surface is nonetheless dominated by its spatially structured part, and we interpret the diagnostics below accordingly.

Global spatial autocorrelation in *b_i_* was assessed with Moran’s I under randomization, using the same Queen contiguity adjacency, row standardized weights, as the model:

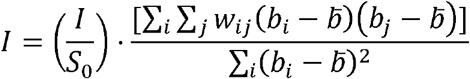

with *S*_0_ the sum over *i* and *j* of the weights *w_ij_*.

Because *b_i_* is itself the output of a spatial smoothing prior, a large positive Moran’s I on *b_i_* is an expected consequence of the model’s own construction and of the estimated mixing parameter φ, and should not be read, as an initial pass through this analysis mistakenly did, as evidence of clustering that the model has failed to capture. The correct reading, adopted here, is that the magnitude of Moran’s I on *b_i_* quantifies how spatially coherent, as opposed to locally erratic, the estimated district level surface is, which is informative about the credibility of the smoothing but is not a residual diagnostic in the sense of testing an assumption against raw data. Local indicators of spatial association were computed as local Moran’s I statistics on the same surface, following the framework of (26), with districts classified into High-High, Low-Low, High-Low and Low-High categories by combining the sign of the local statistic with each district’s position relative to the national mean of *b_i_*, and a district specific pseudo p value threshold of 0.05, uncorrected for multiple testing, consistent with the exploratory, hypothesis generating role of such maps in this context

### 2.7 Model comparison and validation

We report the deviance information criterion, the Watanabe-Akaike information criterion and their associated effective parameter counts, together with the conditional predictive ordinate, a leave one out pseudo marginal likelihood diagnostic computed analytically by INLA without refitting the model. We did not construct an independent held out validation sample, for example by refitting on two survey rounds and predicting the third; the reported diagnostics are therefore internal, in sample measures of fit and complexity rather than external measures of predictive accuracy.

## 3. Results

### 3.1 Crude prevalence and the case for smoothing

Table 1 summarizes unsmoothed national testing coverage. Crude prevalence more than doubled between 2008 and 2014 and continued to rise, though more slowly, through 2022.

The persistence of a substantial minority of very low n districts even in 2022, 11.4% with fewer than 20 respondents, confirms that district level surveillance in Ghana continues to require model based smoothing rather than direct estimation, notwithstanding the expanding GDHS sample size over time and notwithstanding the useful regional picture that single round design based analyses such as (14) have already provided.

### 3.2 Spatial pattern of smoothed prevalence

The fitted surfaces in Figure 1 show a national increase in testing uptake concentrated in the southern half of the country, with the three southern coastal and forest belt regions moving from predominantly under 25% coverage in 2008 to the 50 to 80% range by 2022. A contiguous block of districts spanning the upper middle of the country, corresponding to parts of the Bono, Bono East and Ahafo region cluster, remains visibly darker, lower estimated prevalence, than its surroundings in all three panels, indicating a location specific shortfall that has persisted through fourteen years of otherwise substantial national improvement rather than a transient artefact of any single survey round.

**Figure 1.**
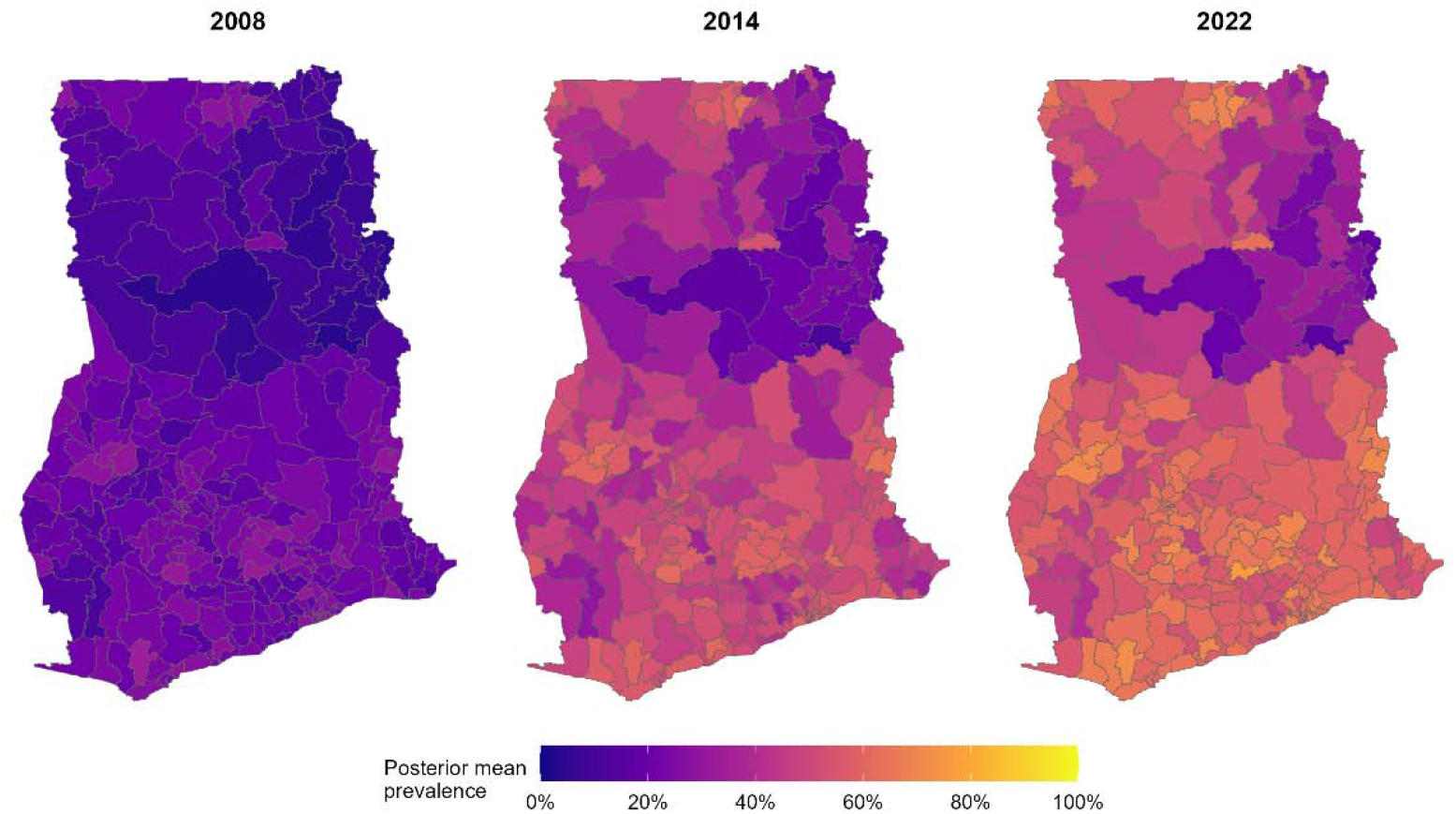
District level posterior mean HIV testing prevalence, GDHS 2008, 2014 and 2022, from the BYM2 and RW1 binomial model.

**Figure 2.**
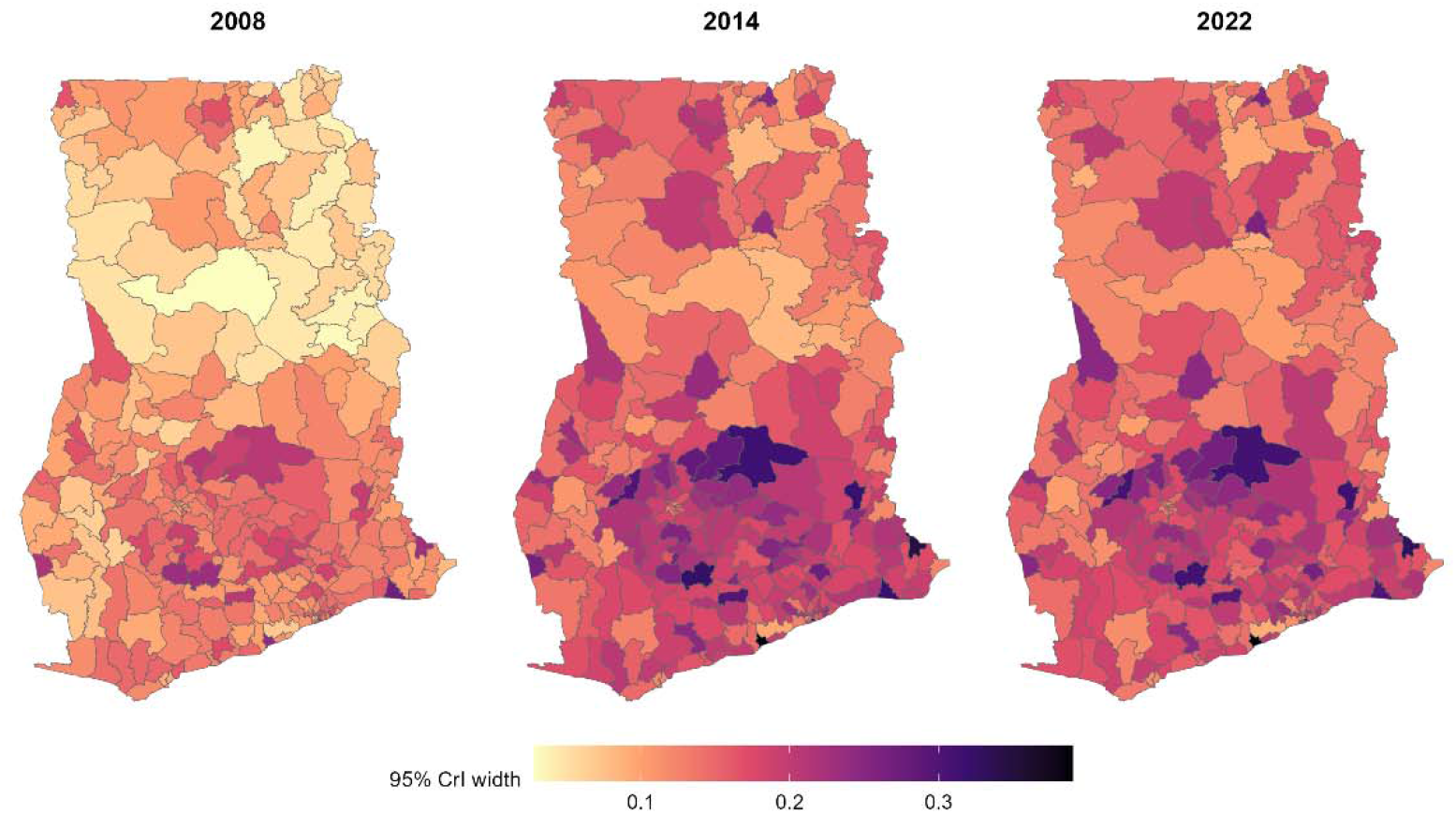
Width of the 95% credible interval of district level posterior mean prevalence, by round. Because interval width on the probability scale compresses automatically as p sub it approaches 0 or 1, binomial variance p(1 - p) is smallest near the boundaries, the apparently narrow intervals in the far north districts in 2008 reflect the very low fitted prevalence there rather than unusually informative data; the corresponding districts have some of the smallest cluster counts in the entire dataset.

**Figure 3.**
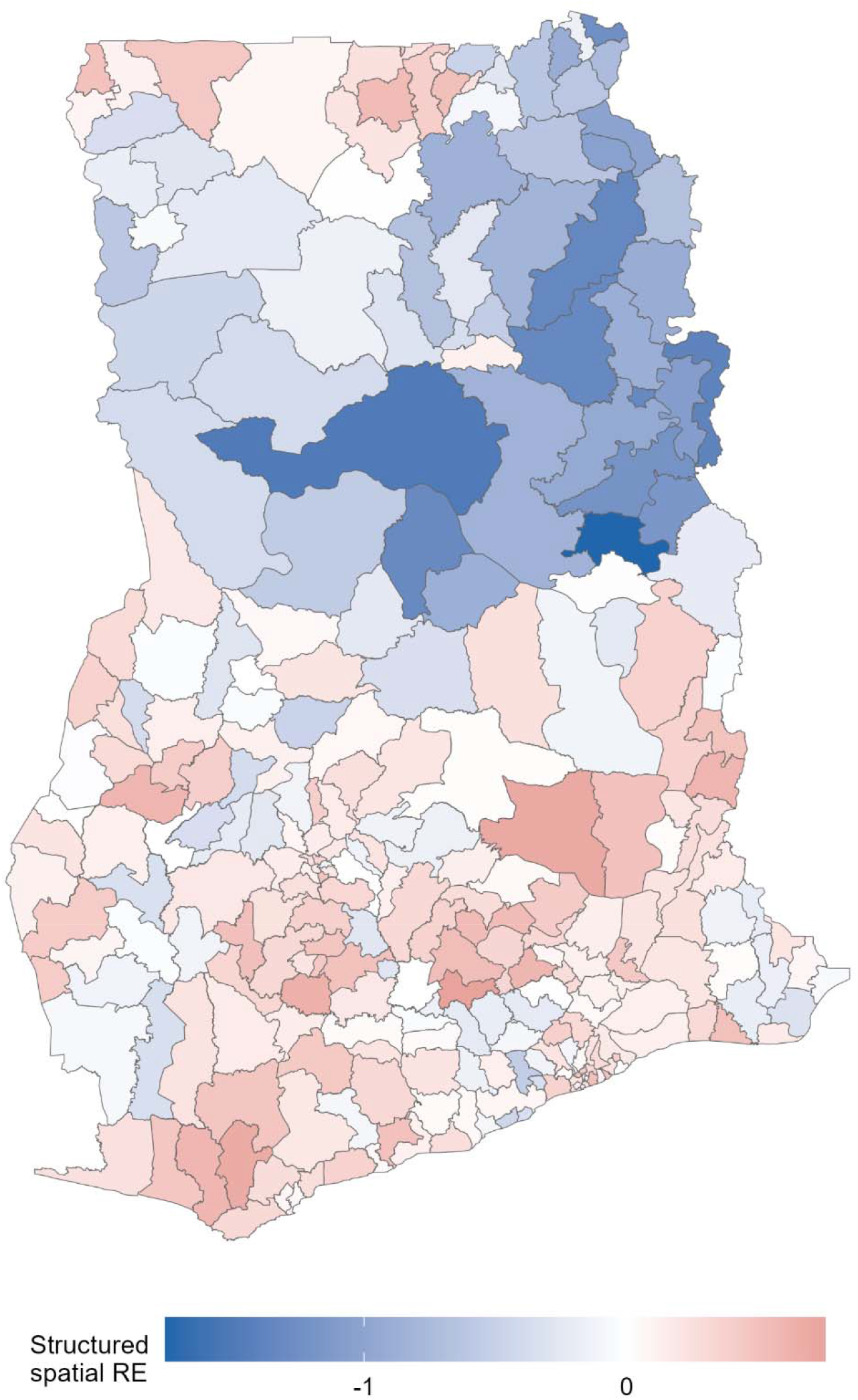
Posterior mean of the combined BYM2 spatial effect. Blue indicates lower than covariate predicted testing uptake; red indicates higher. A contiguous blue band spans the Bono, Bono East and Ahafo corridor in the mid country transition zone; scattered small positive, red, clusters appear in the southern and eastern periphery.

### 3.3 Fixed effects and hyperparameter estimates

**Table 3.** Posterior summaries of fixed effects (logit scale) and corresponding odds ratios.

| Covariate | Posterior mean (logit) | 95% CrI (logit) | Odds ratio | 95% CrI (OR) |
| --- | --- | --- | --- | --- |
| Intercept | -0.452 | -0.497, -0.407 | 0.64 | 0.61, 0.67 |
| WorldPop density (Z) | -0.048 | -0.119, 0.022 | 0.95 | 0.89, 1.02 |
| Travel time to city (Z) | -0.106 | -0.175, -0.038 | 0.90 | 0.84, 0.96 |
| Urban cluster proportion (Z) | 0.097 | 0.043, 0.152 | 1.10 | 1.04, 1.16 |

Urban cluster proportion and travel time were both independently associated with testing uptake in the directions expected from the access to care literature reviewed in Section 1: each one standard deviation increase in a district round’s urban cluster proportion was associated with 10% higher odds of testing, 95% CrI 4 to 16%, and each one standard deviation increase in travel time to the nearest large city with 10% lower odds, 95% CrI 4 to 16% lower. This mirrors the rural to urban gradient reported at the individual level by (3) and by (4), now shown to hold as an independent district level association after accounting for population density and spatial and temporal structure. WorldPop population density carried no detectable independent association once these two covariates were included, with a credible interval spanning unity; given the moderate to high correlation typically observed between raw population density and urban classification, this is most plausibly a case of the urban proportion covariate already capturing the relevant signal, rather than evidence that population density is causally irrelevant to testing access.

**Table 4.** Posterior summaries of variance component hyperparameters.

| Hyperparameter | Posterior mean | 95% CrI |
| --- | --- | --- |
| Precision, BYM2 spatial term ($\tau_b$) | 5.03 | 3.74, 6.59 |
| Mixing proportion, BYM2 ($\phi$) | 0.716 | 0.498, 0.887 |
| Precision, RW1 temporal term ($\tau_\gamma$) | 2.48 | 0.61, 6.54 |

The BYM2 mixing parameter phi = 0.716, 95% CrI 0.498 to 0.887, indicates that the large majority of spatially attributable variance in district level testing uptake is structured, that is shared with geographically neighbouring districts, rather than idiosyncratic to individual districts, even after conditioning on urbanicity, accessibility and population density. The RW1 temporal precision carried a wide credible interval, 0.61 to 6.54, which is unsurprising with only three time points informing a single smoothness parameter, and we caution against over interpreting the shape of the fitted temporal trend beyond the monotonic rise already evident in the crude data in Table 1 and consistent with the trend decomposition reported nationally by (11) over the same 2008 to 2022 window.

### 3.4 Model fit

Deviance information criterion was 3608.6, with an effective number of parameters of 182.6 against 1222.4 under the saturated model; the Watanabe-Akaike information criterion was 3730.7 with an effective parameter count of 244.3; the marginal log likelihood was minus 1783.3. Conditional predictive ordinate based cross validated predictive checks showed no districts with pathologically low values indicative of gross model misfit. These are internal fit statistics; no held out predictive validation was performed.

### 3.5 Residual spatial structure

Global Moran’s I on the fitted spatial effect surface was 0.616, expectation under the null of no spatial structure minus 0.004, variance 0.00145, standardized deviate z = 16.28, p = 6.6 x 10 to the power minus 60. Consistent with the interpretive point made in Section 2.6, we read this as confirmation that the estimated district level surface is strongly spatially coherent, which follows in large part from the estimated phi of 0.716, rather than as evidence of an unmodelled residual pattern that the BYM2 term has somehow failed to absorb.**_=_**

The classification in Figure 4 resolves the global pattern into a single large Low-Low cluster covering essentially the entire northern half of the country, Upper West, Upper East, Northern and northern Bono and Bono East districts, three compact High-High clusters located in the south central Ashanti and Bono-Ahafo corridor and one in the eastern part of the country, and two small High-Low transitional strips lying between the northern low cluster and the southern high clusters, districts whose estimated effect is above the national mean despite being surrounded by below mean neighbours. The continuous local Moran’s I surface, supplementary Figure S1, shows the largest local statistics, exceeding 6, concentrated at the boundary between the northern Low-Low cluster and the adjoining high value corridor, consistent with a genuine step change in testing uptake at that geographic transition rather than a diffuse, gradually varying gradient.

**Figure 4.**
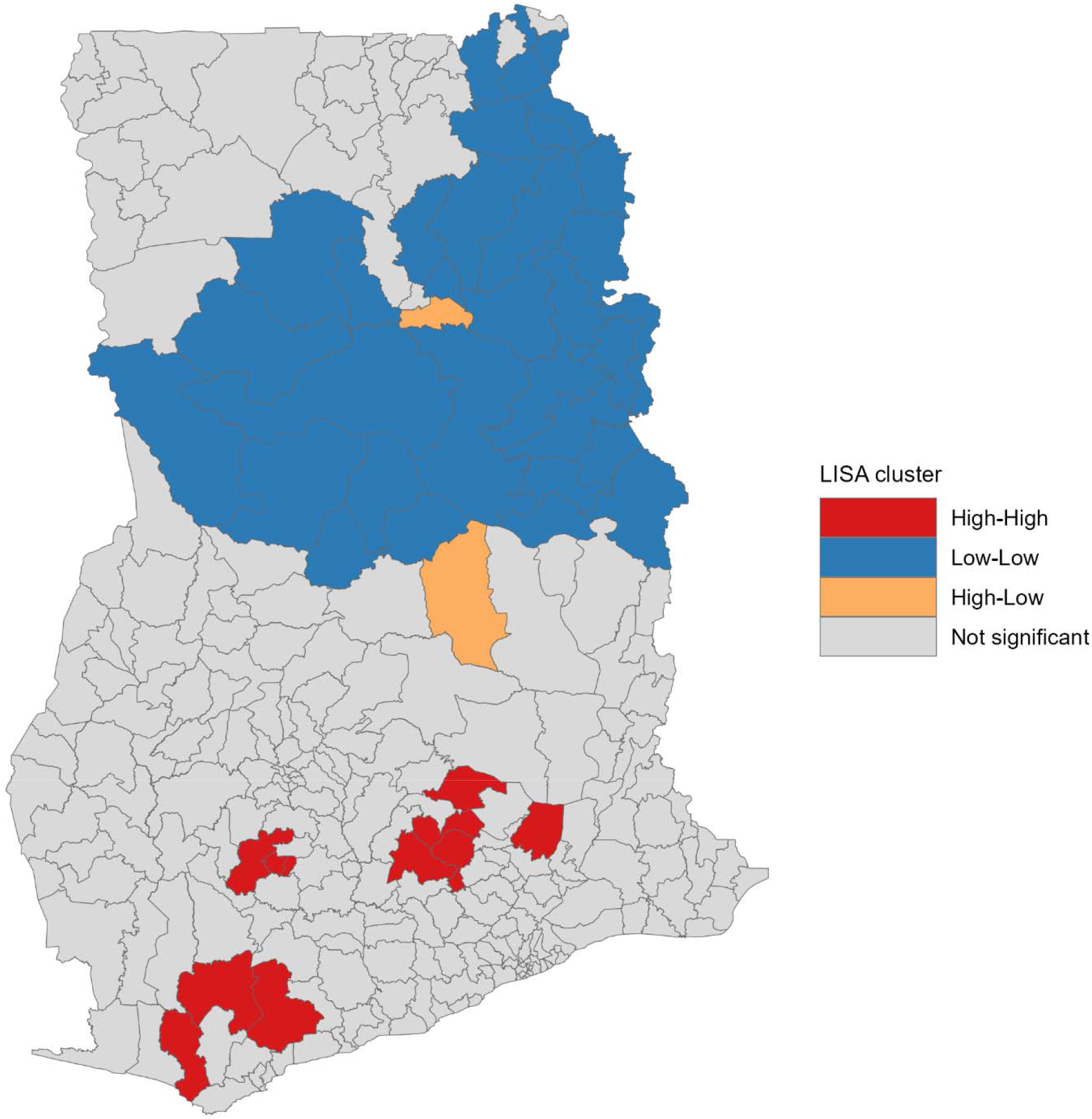
Local indicators of spatial association classification of the district level spatial effect, p < 0.05, uncorrected.

**Supplementary Figure S1.**
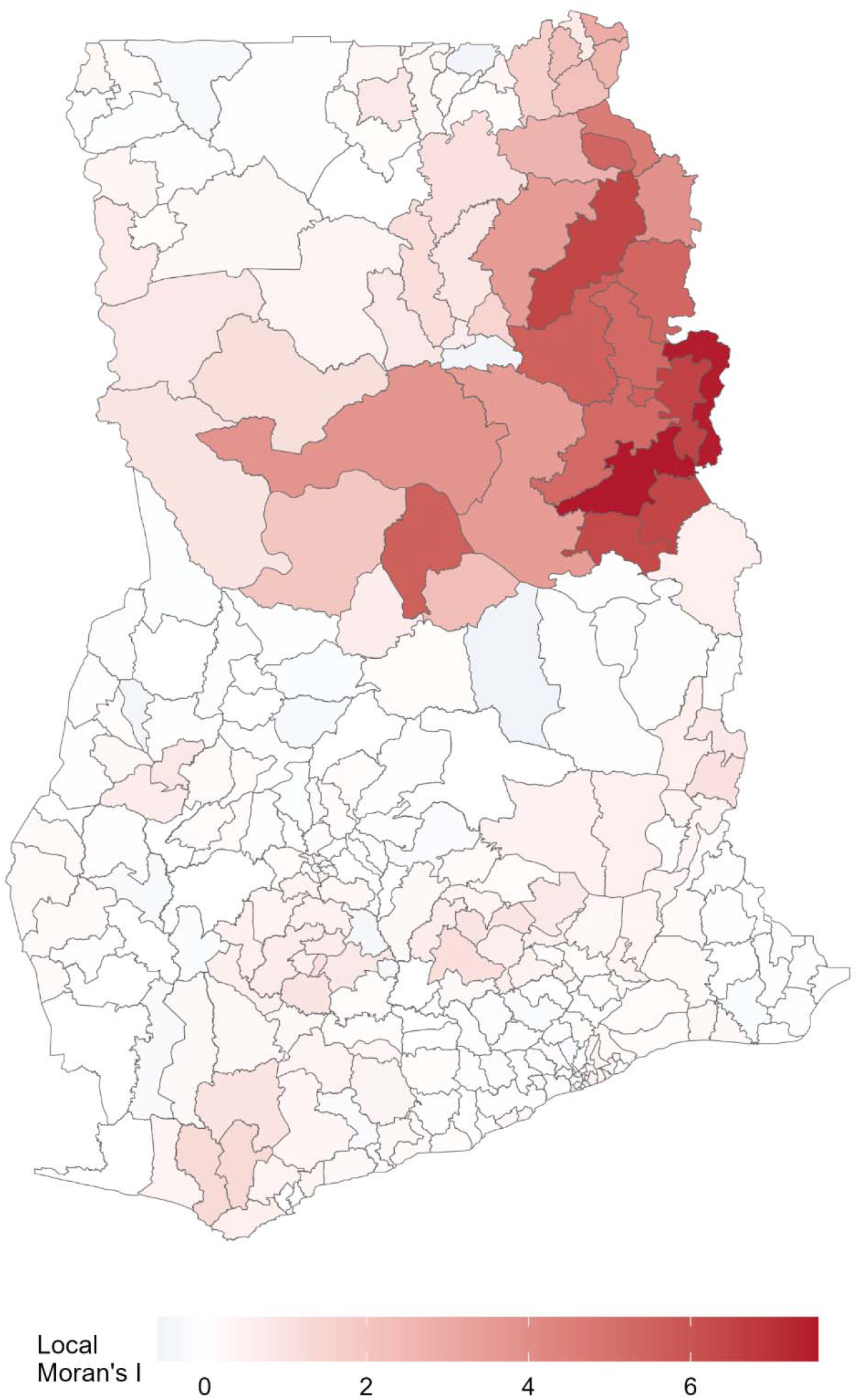
Continuous local Moran’s I statistic for the district level spatial effect.

## 4. Discussion

### 4.1 A persistent north to south gradient beneath a rising national average

The headline national trend, testing prevalence roughly doubling from 2008 to 2014 and continuing to rise through 2022, is consistent with the broader scale up of HIV testing services across sub-Saharan Africa over the same period and with the antenatal testing decomposition reported nationally by (11)across the identical survey window. What the district level model adds to that already documented national trajectory is evidence that the improvement has not closed the geographic gap it began with. The same broad northern corridor that showed the lowest estimated uptake in 2008 remains the lowest uptake region in 2022, both in the smoothed prevalence surfaces in Figure 1 and in the independent LISA classification of the residual spatial effect after adjusting for urbanicity, accessibility and population density in Figure 4. A rising national average has not lifted all districts equally, a pattern also visible at the individual level in the rural to urban disparities reported by (3) and (4), and in the regional access gaps mapped by Ansu-Mensah et al. (2024) for point of care diagnostics in the Bono Region.

### 4.2 Relation to prior small area and survey based work on HIV testing in Ghana

(14) mapped 2014 GDHS testing prevalence across Ghana using design based methods and identified broadly similar regional disparities from a single cross section; the present analysis extends that snapshot into an explicit multi round spatiotemporal model, which is what allows us to distinguish a persistent structural gradient from a transient feature of any one survey round, and to attach formal, propagated uncertainty to every district round estimate rather than only to regional aggregates. The individual level correlates identified across the Ghanaian testing literature, rural residence in (3), health service utilization patterns among men in (1) and (2), sub population specific barriers among students and youth in (6) and (7), and antenatal testing trends in (11), together sketch a picture of who is less likely to test; our district level estimates sketch a complementary picture of where testing uptake remains lowest once those same individual level factors are approximated at the district scale through urban proportion and accessibility. Read together, the two pictures reinforce rather than contradict each other: the districts flagged as Low-Low in Figure 4 are concentrated in the same northern regions where rural residence, lower health facility density and longer travel times, the factors highlighted by (15) and by the national correlates literature, are most prevalent.

### 4.3 What the mixing parameter and Moran’s I jointly imply

An estimated BYM2 mixing parameter of 0.716 means that, of the variance in district level testing uptake attributable to geography at all, roughly seven parts in ten are shared with a district’s immediate neighbours rather than confined to the district itself. Combined with the strongly positive Moran’s I on the fitted spatial surface, this points toward drivers of testing uptake that operate at a scale larger than a single district, plausibly the catchment structure of regional referral hospitals, the reach of specific district health directorates’ community outreach programmes, or historical patterns of donor and non-governmental organization programme placement, rather than drivers confined to any one administrative unit. This has a direct implication for how the LISA clusters in Figure 4 should be read: because the underlying process is spatially contiguous rather than a patchwork of independent district effects, an intervention targeted at the boundary districts of the northern Low-Low cluster is more likely to have spillover benefit for adjacent districts than an intervention of equivalent intensity concentrated in an isolated low uptake district elsewhere.

### 4.4 Methodological contribution

Three aspects of this analysis go beyond the existing Ghana specific literature. First, the explicit RW1 temporal term allows each district’s estimate in a given round to borrow strength from its own history in adjacent rounds, not only from its geographic neighbours in the same round, which the single round design of (14)and the individual level survey analyses reviewed in Section 4.2 could not do by construction. Second, the inclusion of population and accessibility surfaces derived from machine learning methods, standardized and buffered to match the DHS’s own positional uncertainty protocol, supplies exposure information at a spatial resolution the survey sample cannot itself deliver in low cluster density districts. Third, we have tried to state plainly a technical point that is easy to get backwards: a large Moran’s I computed on a spatially smoothed random effect surface confirms that the smoothing has produced a spatially coherent surface, and is a statement about the estimated field’s own construction as much as about the underlying process; it is not, by itself, evidence of a residual pattern the model has failed to capture, and treating it as such would be circular. We think this distinction matters for how results from BYM2 style models are communicated to a non-technical programme facing audience, where the intuitive but incorrect reading is common.

### 4.5 Comparison with small area estimation of HIV indicators elsewhere in sub-Saharan Africa

The pattern we observe, strong spatially structured residual variation even after including plausible covariates, echoes findings from small area estimation of HIV indicators elsewhere on the continent. (18) found comparable spatial dominance in a Bayesian predictive model of HIV prevalence and burden in Nigeria, and (19) reported similarly persistent district level heterogeneity in Zambia after adjusting for available covariates. Continent wide mapping by (20) and the age and sex specific extension by (21) likewise found that geography explained substantial residual variance in HIV prevalence across sub Saharan Africa beyond what standard demographic covariates could account for, and (22) built the Naomi tool specifically because district level HIV indicators in the region could not be estimated reliably between survey rounds without an explicit spatial and temporal smoothing mechanism of the kind used here. (27) and (28) found comparable structural gaps in HIV testing coverage specifically, rather than prevalence, across pooled multi country DHS data, which situates the Ghanaian gradient identified here within a broader regional pattern rather than as an isolated national anomaly.

### 4.6 Limitations

Several limitations bear directly on how these estimates should be used. The RW1 temporal term treats the three GDHS rounds as equally spaced despite true gaps of six and eight years; with only three time points we cannot separately identify a genuine nonlinear trend from an artefact of this approximation, and the temporal component of the model should be read as a device for borrowing strength across rounds rather than as a validated shape for HIV testing growth over calendar time. The night time lights covariate, intended as a third machine learning derived proxy for local economic activity, could not be retrieved for any of the three years because of access restrictions at the source repository and was dropped rather than imputed; its omission may partly explain the modest and non-significant coefficient on population density, since night time lights and population density are often complementary rather than redundant proxies for urbanicity in the literature. DHS coordinate displacement, 2 to 5 km and occasionally 10 km, introduces positional error into every covariate extracted by buffering around cluster coordinates; this error is largest, in relative terms, for small or geographically compact districts, and we cannot rule out some degree of covariate attenuation from this source. We report only internal fit statistics, deviance information criterion, Watanabe-Akaike information criterion and conditional predictive ordinate; no district round was held out and predicted from a model fitted on the remaining data, so claims about the model’s external predictive accuracy, as opposed to its internal fit, are not supported by the analysis as conducted. Finally, the outcome, ever tested for HIV, does not distinguish recent from lifetime testing and cannot on its own monitor the recency of testing indicators used in current cascade monitoring frameworks; the repeat testing behavior documented nationally by (29) among clients already accessing antiretroviral therapy and testing services suggests that a recency weighted outcome, where available, would be a more policy relevant target for a future iteration of this pipeline.

### 4.7 Toward architecturally embedded machine learning

The machine learning contribution in this analysis is confined to the covariate generation stage. (23) demonstrated that a graph neural network can be embedded directly inside a small area estimation framework together with spatial regularization and Bayesian inference, an architectural fusion rather than a covariate level one, and (24) situated this kind of fusion within a broader movement toward artificial intelligence enabled infectious disease modelling. Ghana’s district boundaries were reorganized between the earlier and later GDHS rounds analyzed here, reflected in the differing district counts underlying the GADM layer over time, and an architecture in the spirit of (23) could in principle absorb that boundary instability directly rather than requiring it to be resolved by mapping historical clusters onto a single, fixed contemporary boundary set as we have done. We regard this as the most promising direction for a subsequent, architecturally deeper version of this pipeline, distinct from the upstream covariate integration reported here.

### 4.8 Programmatic implications

For Ghana’s HIV programme, the district clusters identified in Figure 4 offer a concrete, geographically specific starting point for allocating scarce testing outreach resources. The contiguous northern Low-Low cluster is the largest single block of persistently underperforming districts and, given the spatial spillover reasoning in Section 4.3, is a more efficient target for a regionally coordinated mobile or community testing intervention than an equivalent quantity of resources spread across isolated low performing districts elsewhere, a targeting logic consistent with the sub population specific interventions already piloted in Hohoe and Kintampo by (6) and (7) and with the access focused mapping of (15) in the Bono Region. More broadly, the approach itself, combining routinely available survey rounds with open geospatial covariates in a single spatiotemporal Bayesian hierarchy, is directly transferable to the subnational HIV testing and treatment indicators tracked by Ghana’s national AIDS control programme between DHS rounds, and to the analogous small area estimation problems already addressed for HIV burden in Nigeria and Zambia by (18) and (19).

## 5. Conclusion

District level HIV testing uptake in Ghana rose substantially between 2008 and 2022, but a Bayesian spatiotemporal small area model that borrows strength across both geography and survey round, and that incorporates geospatial covariates derived from machine learning methods, reveals a north to south gradient in uptake that measured accessibility and urbanicity do not fully explain and that has persisted throughout the period of national improvement. The magnitude of the estimated spatial mixing parameter and the global and local Moran’s I diagnostics together indicate that this residual structure operates at a scale larger than individual districts, which has direct implications for how testing scale up resources should be geographically coordinated rather than allocated district by district.

## Data Availability

The datasets supporting the findings of this study are available from the DHS Program[](https://dhsprogram.com/data/) after registration and approval of a data request. Analysis code is available from the corresponding author upon reasonable request.

https://dhsprogram.com

https://www.worldpop.org

https://malariaatlas.org

## Declarations

### Ethics approval and consent to participate

This analysis used publicly available, de-identified secondary data from the Ghana Demographic and Health Surveys, obtained with permission from the DHS Program, ICF International. The original surveys received ethical approval from the Ghana Health Service Ethics Review Committee and the ICF International Institutional Review Board, and all respondents provided informed consent at the time of interview. No additional ethical approval was sought for this secondary analysis, consistent with DHS Program data use policy.

### Data availability

Individual recode and geographic datasets for the 2008, 2014 and 2022 Ghana DHS are available from the DHS Program at https://dhsprogram.com upon registration and approval of a data use request. WorldPop population surfaces are freely available at https://www.worldpop.org, and the Malaria Atlas Project accessibility surface at https://malariaatlas.org. Derived district round aggregate datasets and model output tables underlying this analysis are available from the corresponding author on reasonable request.

### Author contributions

Osman Abdul-Fatawu Iddrisu conceived, designed, analyzed and wrote major parts of the manuscript. Hudu Abubakar Siddick performed the data curation, formal analysis, writing, reviewing, and editing. Abubakar Iddrisu Siddiq wrote the methodology, discussion, and performed data quality checks on the spatial covariates. The final draft of the manuscript was reviewed, edited, and revised for intellectual content, read, and approved by all authors.

### AI usage declaration

Claude by Anthropic, was used to assist in drafting and structuring this manuscript from analysis outputs, data tables, model summaries and figures, all of which generated independently by the authors R and INLA analysis pipeline. The authors were responsible for the study conception, data acquisition and processing, all statistical modelling decisions, model structure, prior specification and covariate selection, execution of the analysis, generation of all numerical results and figures. The authors take full responsibility for the content, accuracy and interpretations presented in this manuscript.

## References

1. Seidu AA. Using Anderson’s Model of Health Service Utilization to Assess the Use of HIV Testing Services by Sexually Active Men in Ghana. Front Public Health. 2020;8:512. doi:10.3389/fpubh.2020.00512

2. Nyarko S, Sparks C. Levels and determinants of HIV testing uptake among Ghanaian men. African Journal of AIDS Research. 2020;19:40–7. doi:10.2989/16085906.2019.1679851

3. Dey N, Ansah KO, Norman QA, Manukure JM, Brew ABK, Dey EA, et al. HIV Testing among sexually active Ghanaians: an examination of the rural-urban correlates. AIDS Behav. 2022;26:4063–81. doi:10.1007/s10461-022-03731-4

4. Nketiah-Amponsah E, Ampaw S, Bawuah A. Rural-urban determinants of HIV/AIDS testing uptake among Ghanaian women of reproductive age. BMC Public Health. 2025;25:2783. doi:10.1186/s12889-025-24072-6

5. Iddrisu AK, Opoku-Ameyaw K, Bukari FK, Mahama B, Akooti JJA. HIV Testing Decision and Determining Factors in Ghana. World J AIDS. 2019;9(2):85–104. doi:10.4236/wja.2019.92007

6. Agamlor E, Pencille LB, Lutala PM, Akoku DA, Tarkang EE. Uptake of HIV testing and counseling among tertiary institution students in the Hohoe Municipality, Ghana. J Public Health Afr. 2020;10(2):1044. doi:10.4081/jphia.2019.1044

7. Kabiri M, Akuffo KO, Nortey P, Eusebi P, Danso-Appiah A. Uptake of HIV Testing and Counselling among the Youth in Kintampo South District, Ghana [Internet]. 2024. Available from: 10.60692/jpfx4-sgj30

8. Akweh TY, Boakye B, Adoku E, Teyko F, Tarkang E. HIV testing uptake and its associated factors among Ghanaian men: insights from the 2022 Ghana demographic and health survey using the Anderson behavioral model. Discover Public Health. 2025;22. doi:10.1186/s12982-025-00609-3

9. Akweh TY, Adoku E, Mbiba F, Teyko F, Brinsley TY, Boakye B, et al. Prevalence and factors associated with knowledge of HIV Self-Test kit and HIV self-testing among Ghanaian women: multi-level analyses using the 2022 Ghana demographic and health survey. BMC Public Health. 2025;25. doi:10.1186/s12889-025-21694-8

10. Saaka S, Antabe R, Luginaah I. Correlates of HIV testing among men in Ghana: Cross sectional analysis of the 2022 demographic and health survey. Int J STD AIDS. 2025;36:487–97. doi:10.1177/09564624251324976

11. Alhassan A, Doe P, Salifu Y, Gunu AI, Lasong J, Amoadu M. Trends and inequalities in HIV testing uptake among pregnant women during antenatal care in Ghana: a decomposition analysis from 2008 to 2022. Trop Med Health. 2026;54. doi:10.1186/s41182-025-00897-0

12. Essuman M, Mohammed H, Kebir MS, Obiribea C, Ahinkorah B. Prevalence and factors associated with HIV testing among young women in Ghana. BMC Infect Dis. 2024;24. doi:10.1186/s12879-024-09068-8

13. Boateng R, Boakye D, Kumah E. Predictors of HIV testing uptake among men in Ghana: insights from the 2017/2018 multiple indicator cluster survey. BMC Public Health. 2026;26. doi:10.1186/s12889-026-26279-7

14. Nutor JJ, Duah HO, Duodu PA, Agbadi P, Alhassan RK, Darkwah E. Geographical variations and factors associated with recent HIV testing prevalence in Ghana: spatial mapping and complex survey analyses of the 2014 demographic and health surveys. BMJ Open. 2021;11. doi:10.1136/bmjopen-2020-045458

15. Ansu-Mensah M, Ginindza T, Amponsah SK, Shimbre M, Bawontuo V, Kuupiel D. Geographical Access to Point of care diagnostic tests for diabetes, anaemia, Hepatitis B and human immunodeficiency virus in the Bono Region, Ghana. BMC Health Serv Res. 2024;24. doi:10.1186/s12913-024-11830-2

16. Salifu Y, Walana W, Lasong J, Yakubu M, Wobi EB, Torpey K. Young women’s healthcare screening behaviours and sexual autonomy in Ghana: a spatial distribution and socioeconomic inequality analysis of a large population based survey. Frontiers in Reproductive Health. 2026;8. doi:10.3389/frph.2026.1751165

17. Wakefield J, Okonek T, Pedersen J. Small Area Estimation for Disease Prevalence Mapping. International Statistical Review. 2020;88:398–418. doi:10.1111/insr.12400

18. Onovo A, Others. Estimation of HIV prevalence and burden in Nigeria: a Bayesian predictive modelling study. eClinicalMedicine, 62 [Internet]. 2023. Available from: 10.1016/j.eclinm.2023.102098

19. Mweemba C, Hangoma P, Fwemba I, Mutale W, Masiye F. Estimating district HIV prevalence in Zambia using small area estimation methods. Popul Health Metr. 2021;20. doi:10.1186/s12963-022-00286-3

20. Dwyer-Lindgren L, Others. Mapping HIV prevalence in sub-Saharan Africa between 2000 and 2017. Nature. 2019;570:189–93. doi:10.1038/s41586-019-1200-9

21. Haeuser E, Others. Mapping age and sex specific HIV prevalence in adults in sub-Saharan Africa, 2000 to 2018. BMC Med. 2022;20. doi:10.1186/s12916-022-02639-z

22. Eaton J, Others. Naomi: a new modelling tool for estimating HIV epidemic indicators at the district level in sub Saharan Africa. J Int AIDS Soc. 2021;24. doi:10.1002/jia2.25788

23. Liu P, Chen Y, Liang X, Li H, Biljecki F, Stouffs R. A graph neural network for small area estimation: integrating spatial regularisation, heterogeneous spatial units, and Bayesian inference. International Journal of Geographical Information Science. 2025;40:2358–96. doi:10.1080/13658816.2025.2597971

24. Kraemer M, Others. Artificial Intelligence for Modelling Infectious Disease Epidemics. Nature. 2025;638:623–35. doi:10.1038/s41586-024-08564-w

25. Ngcobo S, Mntla EM, Shock JP, Louw M, Mbonambi L, Serite T, et al. Artificial intelligence for HIV care: a global systematic review of current studies and emerging trends. J Int AIDS Soc. 2025;28. doi:10.1002/jia2.70045

26. Anselin L. Local Indicators of Spatial Association, LISA. Geogr Anal. 1995;27(2):93– 115.

27. Zegeye B, Adjei N, Ahinkorah B, Tesema G, Ameyaw E, Budu E, et al. HIV testing among women of reproductive age in 28 sub-Saharan African countries: a multilevel modelling. Int Health. 2023;15:573–84. doi:10.1093/inthealth/ihad03

28. Sabo KG, Others. Factors influencing HIV testing uptake in Sub-Saharan Africa: a comprehensive multi-level analysis using demographic and health survey data, 2015 to 2022. BMC Infect Dis. 2024;24. doi:10.1186/s12879-024-09695-1

29. Adu-Gyamfi R, Addo SA, Baddoo NA, Kenu E, Ashinyo A, Owusu K, et al. HIV retesting prevalence among clients accessing anti-retroviral therapy and HIV testing services in Ghana. PLoS One. 2025;20. doi:10.1371/journal.pone.0316915

